# Optimal test allocation strategy during the COVID-19 pandemic and beyond

**DOI:** 10.1101/2020.12.09.20246629

**Authors:** Jiacong Du, Lauren J Beesley, Seunggeun Lee, Xiang Zhou, Walter Dempsey, Bhramar Mukherjee

## Abstract

Testing for active SARS-CoV-2 infections is key to controlling the spread of the virus and preventing severe disease. A central public health challenge is defining test allocation strategies in the presence of limited resources. In this paper, we provide a mathematical framework for defining an optimal strategy for allocating viral tests. The framework accounts for imperfect test results, selective testing in certain high-risk patient populations, practical constraints in terms of budget and/or total number of available tests, and the purpose of testing. Our method is not only useful for detecting infected cases, but can also be used for long-time surveillance to monitor for new outbreaks, which will be especially important during ongoing vaccine distribution across the world. In our proposed approach, tests can be allocated across population strata defined by symptom severity and other patient characteristics, allowing the test allocation plan to prioritize higher risk patient populations. We illustrate our framework using historical data from the initial wave of the COVID-19 outbreak in New York City. We extend our proposed method to address the challenge of allocating two different types of tests with different costs and accuracy (for example, the expensive but more accurate RT-PCR test versus the cheap but less accurate rapid antigen test), administered under budget constraints. We show how this latter framework can be useful to reopening of college campuses where university administrators are challenged with finite resources for community surveillance. We provide a R Shiny web application allowing users to explore test allocation strategies across a variety of pandemic scenarios. This work can serve as a useful tool for guiding public health decision-making at a community level and adapting to different stages of an epidemic, and it has broader relevance beyond the COVID-19 outbreak.

## Introduction

The importance of testing for SARS-CoV-2 viral infections has been widely accepted by public health professionals around the world. Identifying infected cases early in their infectious period through large-scale testing efforts can help prevent disease transmission, guide contact tracing and isolation strategies, and contribute to estimation of expected healthcare needs. While immunoglobulin antibody tests can evaluate *past* SARS-CoV-2 viral infections, many public health interventions such as contact tracing are based on detection of *active* infections. Several testing options for detecting an active SARS-CoV-2 viral infection are currently available, and these tests have varying levels of accuracy (as characterized by their sensitivity and specificity) and different barriers to access. Expensive RT-PCR tests are based on nasopharyngeal swabs, whereas less accurate rapid antigen tests require only a saliva sample and are much less expensive [1, 2]. Tests may be administered for many reasons, including diagnostic testing for symptomatic or exposed individuals, population surveillance to detect an outbreak, or enhanced screening in high-risk strata (e.g. essential workers). Testing for active infections is key to controlling the spread of the virus and reducing rates of severe outcomes. A central public health challenge is defining “optimal” test allocation strategies under resource constraints and/or multiple competing test options. The discussion in this paper is broadly applicable to test allocation designs, beyond the COVID-19 pandemic.

One ideal testing strategy for estimating the population infection rate is universal random testing [3], where a large random sample of the entire population is tested. This process is repeated regularly to track the pandemic over time. However, this approach requires conducting an enormous number of tests and is impractical for countries or regions such as the United States with large heterogeneous population and limited number of tests. Several approaches have been suggested for allocating tests under resource constraints. Cleevely *et al*. [4] proposed stratified periodic testing for reducing the effective reproduction rate, where tests are administered at different rates (in terms of test frequency and volume) for patients at different levels of infection risk. Although this approach highlights the importance of stratifying the population and prioritizing testing high-risk groups, the authors did not consider how exactly to distribute the tests across different groups. Another approach that has been suggested for finding cases is pooled/group testing [5, 6, 7, 8, 9], where a single test is applied to merged samples from a group of people. Under pooled testing, the number of required tests is dramatically reduced. Nevertheless, dilution due to combined samples is always one of the major concerns in the pooled/group testing approach [6, 8, 7]. Ely *et al*. [10] described allocation of fixed numbers of multiple different test types with different sensitivities and specificities to populations at high/low risk. Under this approach, tests are allocated by a decision-making process for maximizing the value of the tests, mathematically defined as the sum of the test’s specificity and sensitivity weighted by the loss of the corresponding decision error. This problem setting is similar to ours, but the two objective functions are very different, leading to different interpretation of the results. Ely *et al*.’s approach requires quantification of the relative loss of false negatives and false positives for each individual.

In this paper, we develop a comprehensive mathematical framework illustrated in **Figure 1** for defining an optimal test allocation strategy, accounting for (1) imperfect test results, (2) intensified testing in certain patient populations, (3) resource limitations in terms of budget and/or total number of available tests, and (4) the goal of testing. In our proposed approach, tests are allocated across population strata defined by symptom severity and other patient characteristics (e.g., age, comorbidities, occupation), allowing our test allocation plan to prioritize higher risk patient populations. Since the goal of testing may vary at different points of the pandemic, our proposed objective function provides optimal flexible testing strategies across the key phases of monitoring an ongoing disease surge/outbreak (here, called detecting mode) and long-term surveillance for new outbreaks (called surveillance mode). During detecting mode, our approach allocates tests with the goal of finding as many of thepositive cases as we can to guide contact tracing efforts and isolation interventions. During surveillance mode, our approach allocates tests to give a test positive rate near a target threshold (e.g. 3%). An observed test positive rate exceeding this threshold provides an indicator of rising case counts in the population. Unlike universal random testing procedures and many other existing approaches, neither of the proposed methods aims to directly estimate the disease prevalence in the population. However, we show that control of the test positive rate at a target level implies an upper bound on the population disease rate under certain conditions (supplementary materials). Our framework assumes that the true disease status is independent of patient characteristics, and selection is independent of the disease status, conditional on the symptoms and patient characteristics. Furthermore, we assume that the diagnostic test result is independent of other factors (including symptoms) conditional on the true disease status. We also assume that people with severe symptoms are always tested. Defining population strata based on symptoms and age, we use extensive simulation studies to evaluate the proposed method for optimal test allocation strategy based on the purpose of testing. We illustrate how these methods can be applied to determine test allocation through different stages of the pandemic in New York City (NYC).

We also extend this framework to address the question of how to allocate two *different types* of tests with differing accuracy, e.g. RT-PCR tests versus rapid antigen tests. This approach can provide guidance about allocation of multiple types of tests at the local level (e.g., for colleges and universities) under budget constraints. We explore the properties of this approach through simulation.

**Figure 1:**
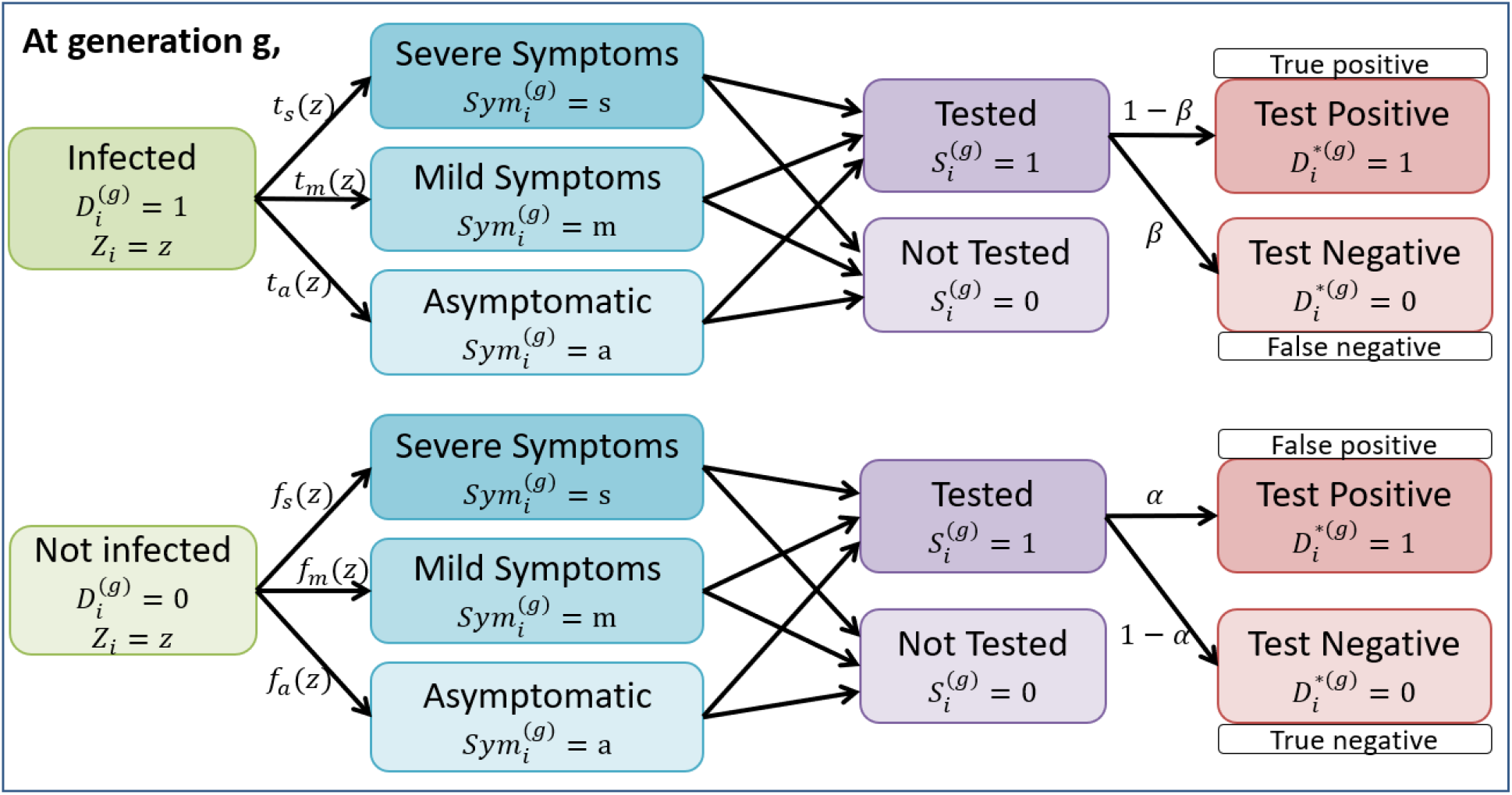
Illustration of data structure at generation *g* with the incorporation of selection and misclassification. For a person *i* with the characteristic information(or risk factors) *Z*_*i*_ *= z* at the beginning of generation *g*, the true disease status 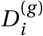 is unobserved, with 1 indicating being infected and 0 being uninfected. The probability for an infected person of developing severe(*s*)/mild(*m*)/no(*a*) symptoms is *t* _*j*_ (*z*), where *j* ∈ {*s, m, a*}, which is based on the characteristics. An uninfected person may also develop similar symptoms due to other diseases, e.g. influenza, and the probability is *f* _*j*_ (*z*). Often, people are tested/selected based on symptoms and some other risk factors. *β* and *α* are the false negative and positive rate for the test.

We provide a R shiny app available at https://umich-biostatistics.shinyapps.io/Testing_Optimization/ that implements all of the proposed methods. In this app, users can specify their goals of testing as well as other key variables to obtain a customized optimal test allocation strategy.

## Results

### Simulations

We explore by simulation how the optimal strategy for allocating a single type of test varies by 1) the number of available tests; and 2) the marginal probability of being asymptomatic for a truly infected individual, 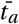. We consider a hypothetical region of population 8 million similar to New York City. Although the true proportion of asymptomatic infections is likely to be unknown, existing literature suggests this proportion varies across regions, with estimates from around 30% to almost 90% [11, 12, 13]. We consider settings with high and low disease prevalence separately, which correspond to being *near* or *past* the pandemic peak, respectively. Other parameter settings can be found in **Methods**.

**Figure 2 a & b** corresponds to the setting with limited number of tests and high population disease rate. In this setting, the majority of tests are allocated to people with severe and mild symptoms unless the probability of being asymptomatic among infected individuals is very high (e.g. *>*0.75). This is because, unless 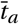 is very large, the probability of a random person with mild symptoms testing positive is larger than the probability of a random person without symptoms testing positive. In contrast, when we have an abundance of tests available (**Figure 2 c & d**), the majority of the tests are allocated to asymptomatic patients, partially due to a limited number of patients with severe and mild symptoms overall. When tests are scarce and the goal is to detect more cases, tests are allocated across all four age groups based on the assumed marginal proportion of infected people who are asymptomatic, 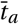. These age distributions among the tested patients are largely driven by differences in symptoms across age categories. Since younger people are more likely to be asymptomatic [11], the majority of tests among asymptomatic people are allocated to young (ages 0-17) people. For example, when the marginal probability of being asymptomatic for an infected person is 0.55 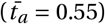 and when we have an abundance of tests as 200, 000, 4.3% of tests are allocated to people aged 65+ with mild symptoms compared to 1.5% to people aged 0-17 with mild symptoms. After satisfying the prioritized testing for the severe and mild symptom groups, the remaining 59.5% of tests are allocated to the young (age 0-17) asymptomatic group.

**Figure 2:**
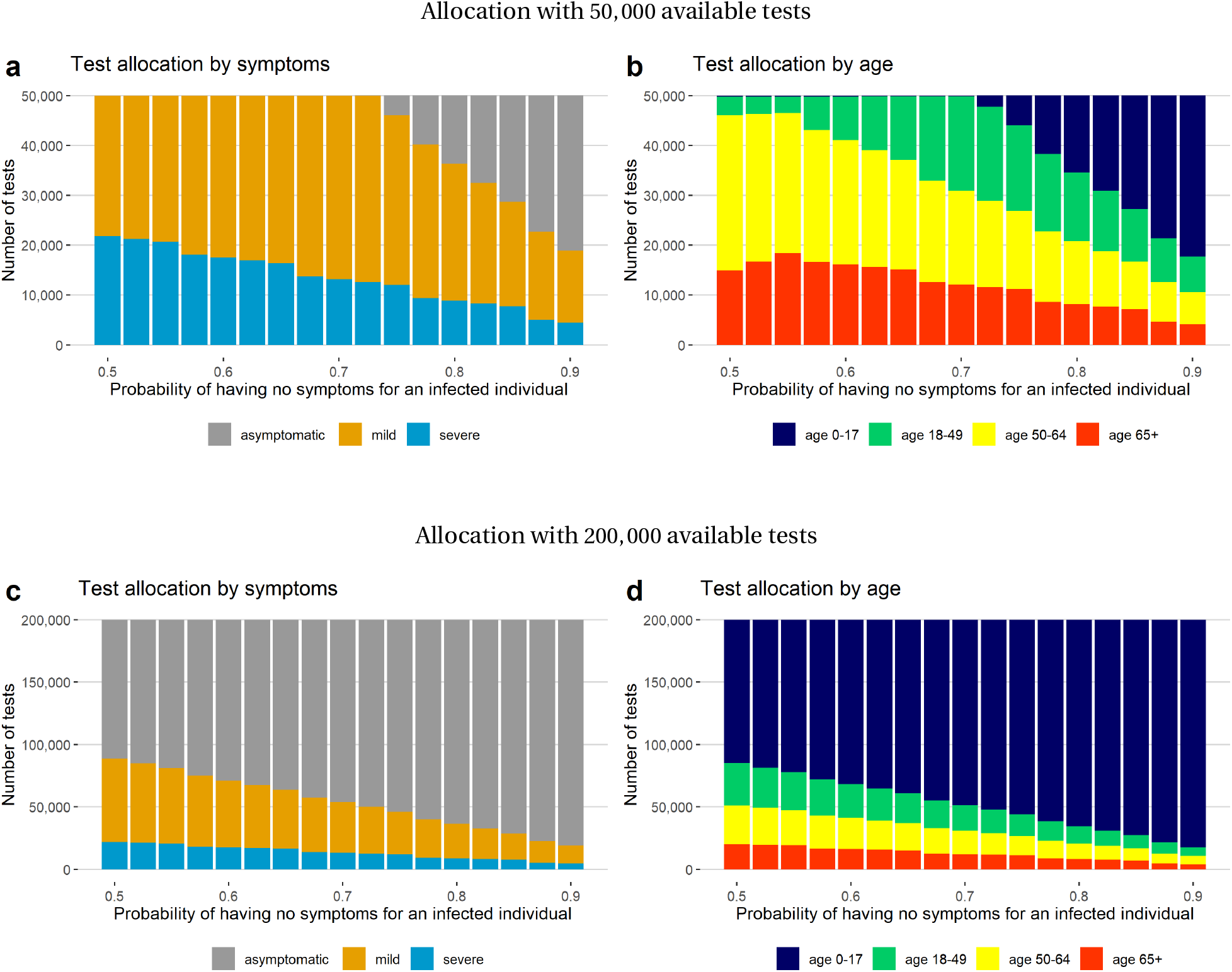
Tests allocated to each symptom and age group near the peak of the pandemic under the detecting mode, assuming either 50, 000 (a) or 200, 000 (b) tests are available and that the number of true infected cases is 175,000 in a region of 8 million people.

**Figure 3** shows the optimal test allocation when the disease prevalence in the population is low and many tests are available. In this situation, we are in a surveillance mode and want to monitor when the test positive rate exceeds a certain threshold (*c*), e.g. 3%. A test positive rate exceeding the threshold level *c* obtained under this testing strategy may provide a good indicator that the prevalence of the disease is going up in the population (**Figure S1** in the supplementary materials). The optimal testing strategy does not require all available tests to be used, and the majority of allocated tests are given to people aged 50+. Close monitoring of older asymptomatic patients may provide a good strategy for capturing an outbreak as indicated by a raising test positive rate.

We compare our proposed optimal strategy to four alternative strategies, denoted as the *risk-based strategy*, the *symptom-based strategy*, the *severe-only strategy*, and the *universal random testing strategy* (**Table S1-S2**). The *risk-based strategy* prioritizes the group with higher risk of being hospitalized; that is, it prioritizes the severe and mild symptomatic people, but within each symptom group, elderly people are always tested first. The *symptom-based strategy* allocates tests based only on the severity of symptoms, and tests are randomly assigned to individuals within symptom groups, regardless of age or other risk factors. The *severe-only strategy* prioritizes testing the severely-ill patients and randomly assigns the remaining tests to the rest of the population. The *universal random testing strategy* randomly tests the entire population without prioritizing any selected group.

**Figure 3:**
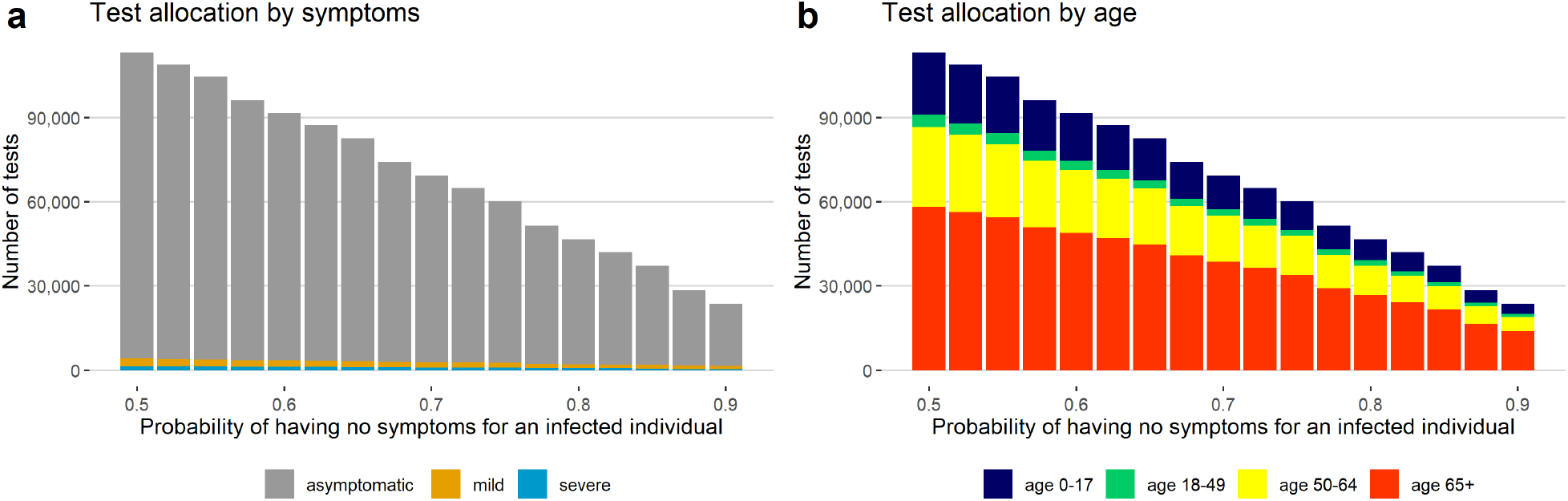
Tests allocated to each symptom and age group for surveillance past the peak of the pandemic, assuming 200, 000 tests are available and the number of true infected cases is low (10,000) in a region of 8 million people. The test positive rate for the disease outbreak is 0.03.

Our proposed optimal strategy under the **detection mode** can identify more cases than all the other methods. In a limited testing resources scenario with 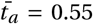, our strategy finds 26 times more cases than the universal random testing. Although the number of identified positive tests from the risk-based strategy is close to ours, these two strategies differ in prioritizing groups of people for testing. Our method prioritizes testing for people aged 50*−*64 over people aged 65*+* among people with mild symptoms, while the risk-based strategy recommends the opposite. The difference between these two test strategies is more apparent when we have an abundance of tests. In this setting, our proposed strategy assigns 59.5% of available tests to the young (aged 0*−*17) asymptomatic people, rather than to the elderly (aged 65*+*) asymptomatic people as the risk-based strategy does. With a sufficient number of tests, our proposed strategy identifies more positive cases than any other method, finding 2.6% more positive tests than the risk-based strategy and 11 times more cases than universal random testing. Our method outperforms all other approaches considered when the probability of being asymptomatic among true cases is higher, e.g.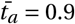.

Unlike the other testing strategies that use all available tests, the proposed optimal test strategy uses only 51.5% of available tests when our goal is conducting **disease surveillance**. For comparison, we also present optimal test allocations obtained from our method under the detection model. Our surveillance model approach finds about 40% fewer positive cases than the proposed method under detecting mode and than purely risk-based and symptom-based strategies, but it still finds more cases than the severe-only and universal random testing strategies. Even using fewer tests than the other methods, our approach can obtain a nominal 3% test positive rate, and observed deviations from this target positive rate can be used as an indicator that the population prevalence is larger than expected.

### A sample case-study: test allocation strategy in New York City

We illustrate how our framework can be applied to determine test allocation through different stages of the pandemic in New York City (*N =* 8, 175, 133) between March 3 and November 1. The case numbers and the total available tests for each week/generation are obtained from data released by the New York City Department of Health and Mental Hygiene [14]. Existing work studying the magnitude of under-reported cases and the proportion of asymptomatic cases suggest that the true number of cases is about 10 times the reported cases in the United States [15]. However, since New York City has tested nearly 70% of its population until November 11 [14], which is far above the national level of 45% [16], the fraction of under-reported cases should be smaller. We assume the multiplicative under-reporting factor to be 4, meaning we assume the true number of cases in the New York City to be 4 times the reported cases. Following Rahmandad *et al*. [15], we set the marginal probability that a true case is asymptomatic, 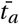, to be 55%. Although the accuracy varies across different types of diagnostic tests, the false negative rate is known to be appreciable and depends on the type of test [17]. We set the false positive (*α*) and false negative (*β*) rates to 0.01 and 0.3 [17], respectively. Other details regarding assumed parameter settings can be found in **Methods** and **Table S3** in the supplementary materials. Alternative parameter values can be explored dynamically using our R shiny app.

**Figure 4** shows the optimal test allocation strategy our method would have recommended for New York City throughout the pandemic. We suppose we had allocated tests with the goal of detecting as many cases as possible between March 3 and July 21, during which the disease prevalence was high in the population. Under this method, we predict that the test positive rate would have fallen below 0.03 during the week of July 21st. We then switched to the surveillance mode for monitoring disease outbreaks thereafter. When the goal is to detect as many cases as possible (detection mode), symptomatic and the elderly patients should be prioritized, especially when we are short of tests. From March 3 to April 7, for example, our method would test only people with severe and mild symptoms, and people of age 0-17 would be rarely tested, because the probability of finding a positive test in the symptomatic group is higher than the asymptomatic group. As the number of available tests gradually increased, more tests would have been allocated to asymptomatic patients and to young people, because the younger people would be more likely to be asymptomatic and the symptomatic elderly people would have already been offered a test. After switching to surveillance mode, our method would have allocated just 53.9% of the tests that are actually conducted. These tests would primarily be allocated to older asymptomatic people.

**Figure 4:**
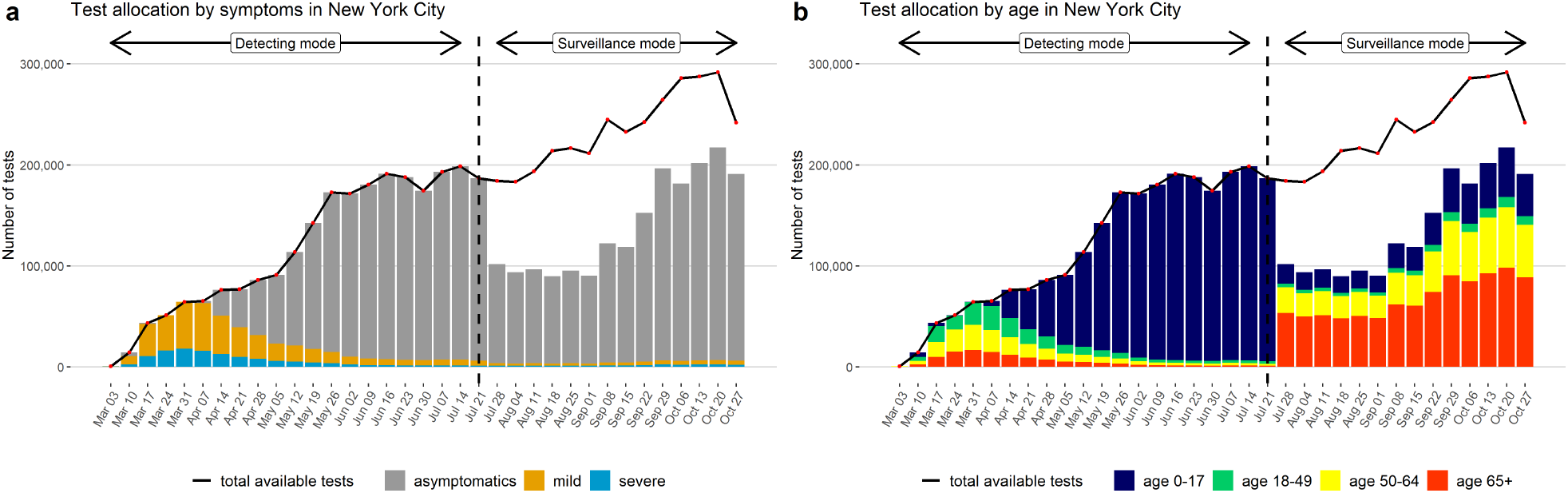
Test allocation strategy for the New York City assuming a case under-reporting factor of 4, stratified by symptoms and age.

To validate our proposed method, we compare the number of detected cases and the test positive rateobservedfor New York City tothepredictedvalues underouroptimaltesting strategy methodin **Figure 5**. Under our assumptions about the rate of case under-reporting, the proposed testing strategy is able to detect a greater number of cases than were actually observed in New York City between March 3rd to July 21st. After July 21st, the optimal test strategy detects a similar number of reported cases but uses far fewer tests than were actually administered during this time period for New York City.

**Figure 5:**
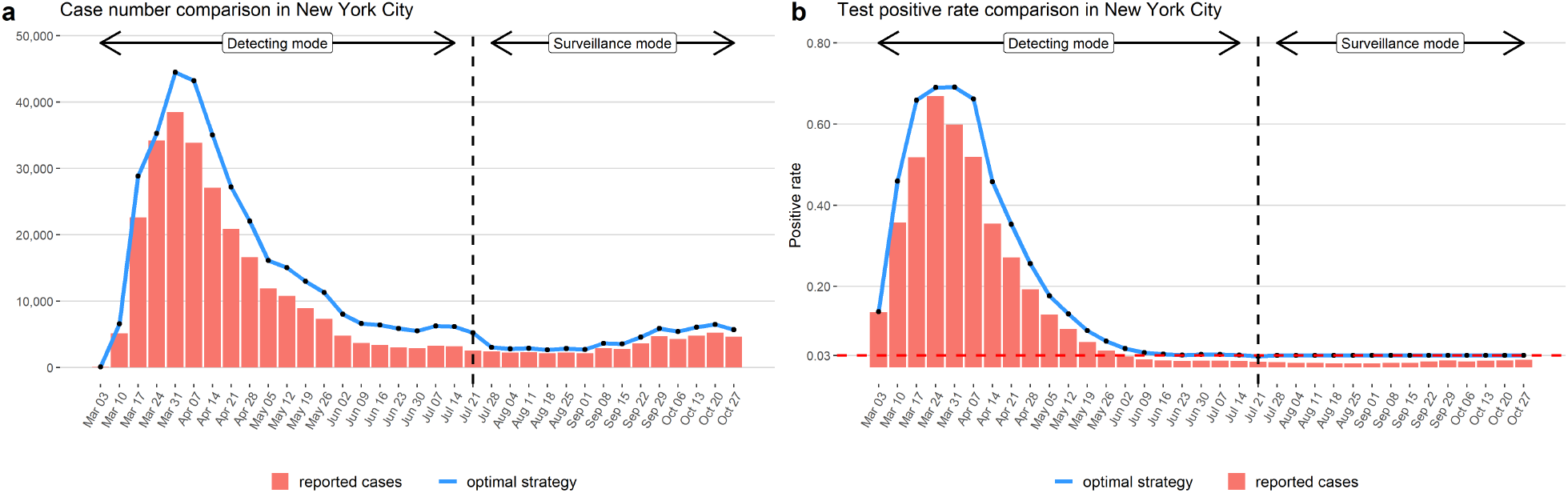
Comparison on predicted case numbers and test positive rates under optimal testing strategy to values observed for New York City assuming a case under-reporting factor of 4.

### Extension to allocating two tests

For universities and colleges, test allocation needs to be customized according to the local disease rates and community testing capacity [18], as are often characterized by the budget for testing, the total number of tests available, the observed number of positive tests and test positive rate (**Table S4**). As tests of different costs and accuracy become available, the question of test allocation becomes even more challenging. Subject to budget constraints, we extend our proposed test allocation method (see **Methods**) to address allocation of two competing tests with differing cost and accuracy as a function of symptoms and patient characteristics. We consider the hypothetical scenario where we want to allocate a fixed budget to a mixture of rapid antigen tests and RT-PCR tests. The RT-PCR test, which is considered to be the gold-standard test in COVID-19 diagnosis, costs about $100 per test and has sensitivity as high as 0.9 [19, 20, 21]. Rapid antigen tests, in contrast, cost as little as $ 5 per test but have much lower sensitivity [19, 20, 22]. Mirroring current market information, we set the price for the rapid antigen test and for the RT-PCR test as $5 and $135, respectively, and we assume test sensitivity values are 0.45 and 0.9, respectively [22, 21]. Specificity is set to 0.99 for both tests. We suppose that at a certain generation, the number of truly infected cases in a population of 8 million is 10,000 and the budget is 1 million. Alternative scenarios (including different budgets, population size, age distributions, etc.) can be explored using our R Shiny app.

**Figure 6** provides the optimal budget allocation between the two tests as a function of the marginal probability thatatruecaseis asymptomatic (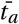). As seeninsupplementary **Figure S2**, RT-PCR testsareonly allocated to people with severe and mild symptoms, and the majority of rapid antigen tests are allocated to asymptomatic people. Since the absolute number of symptomatic people decreases when (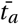) increases under a *fixed number of total cases*, the proportion of the budget allocated to RT-PCR testing decreases with increased (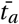).

**Figure 6:**
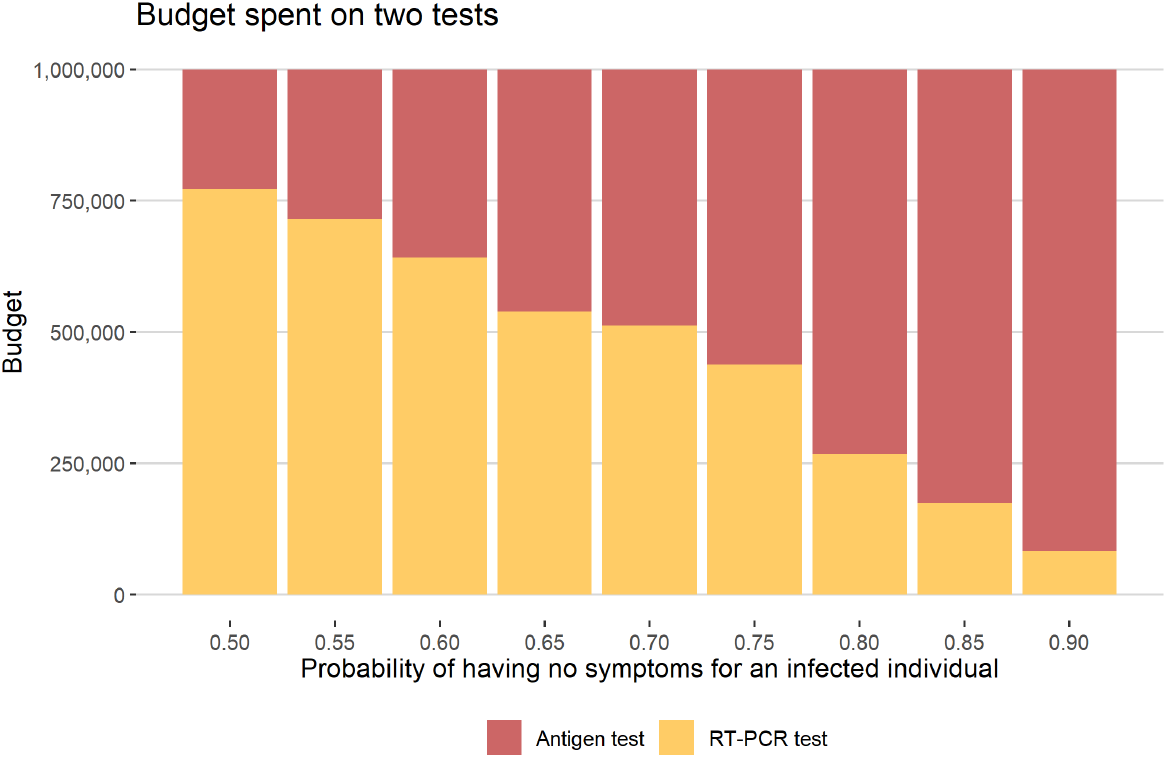
A total of 1 million budge divided for rapid antigen tests and RT-PCR tests. The price of a single antigen test and RT-PCR test are $ 5 and $ 135, respectively. The number of infected cases is assumed to be 10, 000 in a region of population 8 million.

## Discussion

In this paper, we provide a mathematical framework for relating population COVID-19 infection rates to test positive rates as a function of targeted diagnostic testing with imperfect accuracy. We develop a strategy for obtaining optimal allocation of diagnostic tests across population strata defined by symptom severity and other risk factors such as age. This method adapts to different scenarios in terms of public health objectives. For example, when the goal is to detect as many infected cases as possible in the acceleration phase of the pandemic with high community prevalence, tests should be allocated with targeted testing in population strata most likely to contain cases. When our goal is to detect new disease outbreaks as part of population surveillance after a disease wave has passed and we are in a state of containment, fewer tests may be needed, and a substantial proportion of tests should be allocated to asymptomatic people. In the setting where we have a sufficient number of tests, we demonstrate that our proposed detecting mode strategy can find 2.6% more positive tests than the risk-based strategy and 11 times more than universal random testing. Under the surveillance mode, our strategy only uses 51.5% of tests that are available. Although our model assumes that a person’s disease status is independent of his/her characteristics, this assumption can be relaxed if the distribution of true disease status given patient characteristics is known.

We demonstrate this optimal test allocation strategy in a special case where the population is stratified based on age and the severity of symptoms, using New York City as a illustrative example. If tests had been allocated as suggested by our method, we may have used only 53.9% of the tests that were actually conducted and have still found as many cases as were reported. We provide a R Shiny web app (available at https://umich-biostatistics.shinyapps.io/Testing_Optimization/) allowing users to explore the optimal test allocation as a function of test positive/negative rates, number of available tests, and the true rate of infection among asymptomatic people.

In an extension of the proposed method, we develop a strategy for obtaining optimal allocation of two tests with different false negative rates and cost (e.g. cheap rapid antigen tests vs. expensive RT-PCR tests) subject to overall budget constraints. We show that the expensive but more accurate RT-PCR tests should be used on the severe or mild symptomatic people, which is in accordance with the finding in [10]. This approach can be used to help inform test allocation decisions currently being made by many universities, communities, businesses, etc. for planning their reopening. Through our R Shiny app, users can explore the impact of comparative cost, total budget, population age profile, and other key factors on the optimal test allocation at different points in a pandemic wave.

Our R shiny web app can also be used to explore the problem of repeated testing, where rapid antigen tests are repeatedly used to improve sensitivity. We found that the probability of correctly identifying an infected individual goes up with repeated rapid antigen testing, but the ability to detect cases in the entire population decreases as fewer different people are able to be tested under a fixed overall test budget.

An advantage of the proposed test allocation method is that it directly incorporates test accuracy and can be applied to allocation of different types of tests, including fast antigen tests and/or RT-PCR tests. We focus on the particular case where population strata are defined based on age as well as symptom severity, but this method can be extended to also incorporate occupation, geographical location, and other key factors into defining population strata. We provide an example R script for implementing the methods with strata defined by age and symptoms in the supplementary materials. Users can adapt this code to apply our methods under different strata definitions. Care must be taken when specifying model parameters, since the resulting test allocation may be sensitive to these choices. We recommend, therefore, that users explore test allocation across a spectrum of plausible input parameters to inform decision-making in practice.

As the world is planning to disseminate the COVID-19 vaccine [23, 24, 25], inoculation of half of the world will take significant time and will reach different parts of the world at different speeds. Until global herd immunity is reached, testing will be one of the key strategies to manage and contain the disease in many parts of the world. We hope this general framework leads to cost-saving and effective strategies, particularly in developing countries where resources are limited.

## Methods

### Conceptual Framework

Consider a population of size *N*, and let 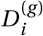 be a binary variable representing person *i* ‘s true (unobserved) disease status (infected-1 vs. not infected-0) at the *g* -th generation of disease circulation. Here, a generation is defined as the average time it takes an infected person to become infectious, which is around 5 days [26]. At the *g* -th generation, some subset of the population will be tested, with binary 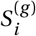 representing whether person *i* in the population is tested during generation *g*. Let 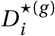 denote the test result (positive-1 vs. negative-0) for person *i* during generation *g*, which may or may not be the correct result (may or may not equal 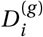). If person *i* is not tested, 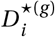 will not be recorded. Let *T* ^(*g*)^ denote the number of tests available for generation *g*. With limited testing capacity, current test strategies prioritize tests based (at least in part) on severity of symptoms. Let 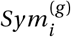 be a categorical variable which takes value in {*s, m, a*} corresponding to severe, mild, and asymptomatic symptom levels respectively. We suppose testing may also depend on other covariates, *Z*_*i*_, such as patient age and occupation.

In modeling symptoms, we assume the probability 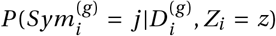 is the same for any *g*. We let *t* _*j*_ (*Z*_*i*_) be the probability of developing the *j* -th symptom conditional on 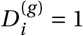 and *Z*_*i*_, and we let *f*_*j*_(*Z*_*i*_) be the probability conditional on 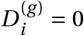. The marginal probabilities of developing symptom *j* in the infected and uninfected population are denoted as 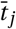 and 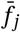. We further make the following three independence assumptions about testing and test results:

A1. 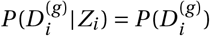,so 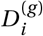 is independent of *Z*_*i*_

A2. 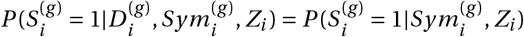, so 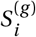 is independent of 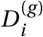 given 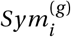 and *Z*_*i*_

A3. 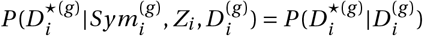, so 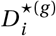 is independent of 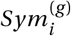 and *Z*_*i*_ given 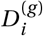.

These assumptions result in the following model structure at generation *g* :

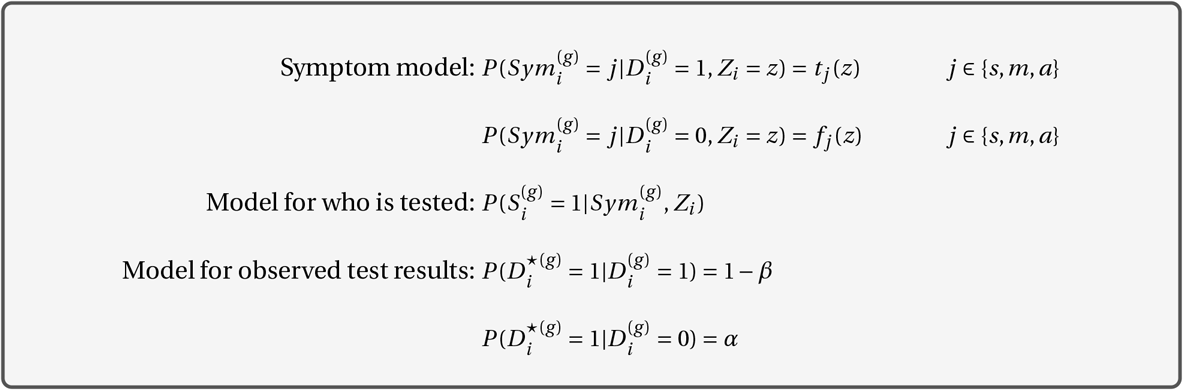

Under this model structure, we can establish the probabilistic relationship between the detection of a positive case and the testing procedure. We use this relationship to predict the number of people testing positive at generation *g*, denoted *P* ^(*g*)^, as a function of test allocation.

### Predicting P ^(g)^, the number of people who will test positive

For a tested/selected person *i*, we express the probability of testing positive as

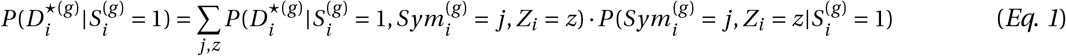

For simplicity, we assume *Z*_*i*_ takes discrete values, and the summation over *Z*_*i*_ in *Eq. 1* would become an integral if *Z*_*i*_ were continuous. The first term of the summation captures the likelihood of a positive test in the selected population given symptoms 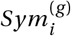 and other covariates *Z*_*i*_. The second term describes the joint distribution of symptoms and *Z*_*i*_ in the selected population. We now take a closer look at each of the terms in the summation.

The probability of person *i* testing positive can be expressed as a function of symptoms 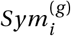 and covariate *Z*_*i*_ as follows:

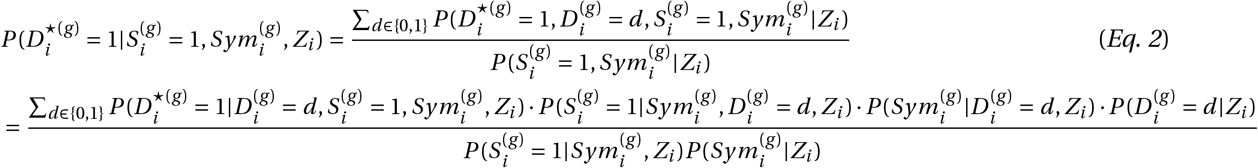

Under assumptions A1-A3, then *Eq. 2* can be written as:

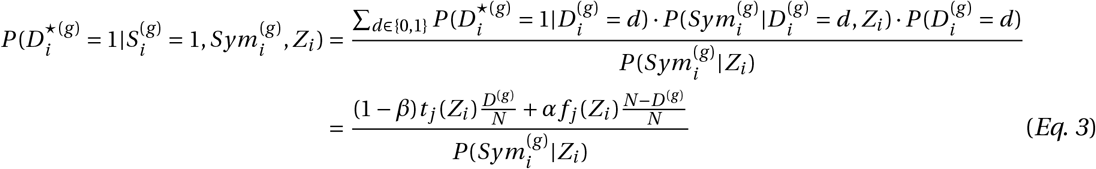

where 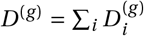 is the number of infected people in the population. The joint distribution of 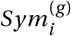 and *Z*_*i*_ for tested people can be expressed as:

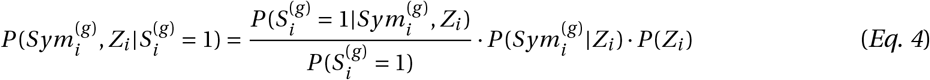

Putting these pieces together, the probability that person *i* in the selected population has a positive test can be expressed as:

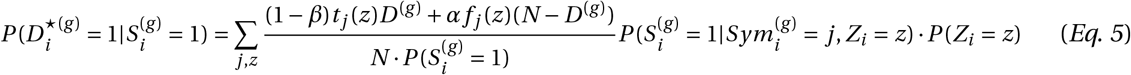

Summing over tested people, the number of positive tests is predicted as

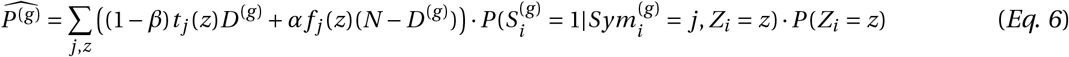

*Eq. 6* depends on 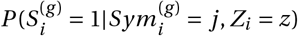, which is the probability of testing in generation *g* for a person *i* with symptoms 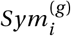 and covariates *Z*_*i*_ *= z*. This term represents the testing protocol in the population, and constraints on testing in terms of (1) test availability and (2) testing prioritization correspond to constraints on 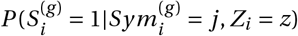.

### Optimal test strategy

Many researchers have been studying the true prevalence of COVID-19 in the population given the number of positive tests [27, 15]. Let *D*^(*g*)^ represent the true number of cases in the population at generation *g*. We then estimate *t* _*j*_ (*z*) and *f* _*j*_ (*z*), the probability of developing symptom *j* given *Z*_*i*_ *= z* and 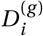 using the historical data (see **Methods Parameter estimates and setup**). Under the above mathematical framework, the problem of test allocation is nothing but to find 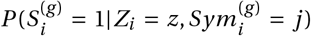. We then construct the following objective function:

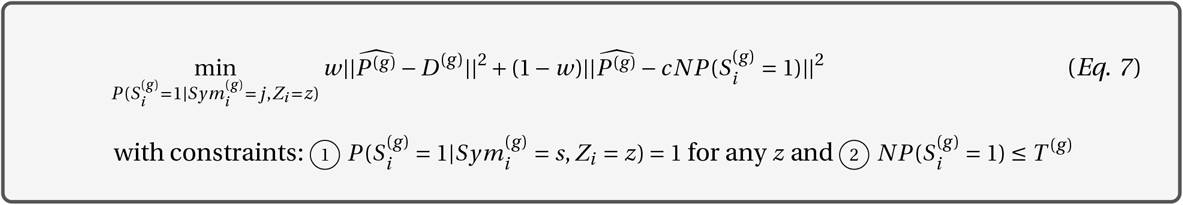

where *c* ∈ (0, 1) is a pre-fixed target test positive rate for detecting the outbreak of the pandemic. The first term 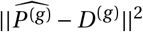 controls the difference between the number of positive tests and the true case counts. The second term 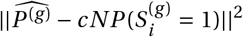 controls the difference between positive rate and the target outbreak threshold *c. w* takes value 0 or 1, indicating the preference for either component in defining the optimal testing strategy. For example, when *w =* 1, the objective function reduces to 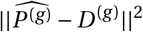, which corresponds to a goal of finding the most accurate test allocation strategy to detect the greatest number of positive tests (detecting mode). On the other hand, *w =* 0 corresponds to a goal of detecting if the test positive rate has crossed a pre-designated outbreak threshold (surveillance mode). In **supplementary materials**, we show that the population disease prevalence is bounded by a function of the test positive rate under mild conditions, so a low test positive rate implies a low disease prevalence in the population. The first constraint in *Eq. 7* ensures that everyone with severe symptoms is prioritized for testing. With limited testing resources, the second constraint guarantees that the number of people tested does not exceed the total number of available tests. We obtain the optimal testing strategy in *Eq. 7* using R package *optiSolve* [28].

Our objective function involves the most commonly-used information metrics for COVID-19, including the overall number of positive tests and the overall test positive rate, which are straightforward and easy to understand for the general public. Neither the detecting mode or the surveillance mode in our objective function aims for estimating the true prevalence in the population, which is usually the goal of universal random testing. It is worthy to mention that if testing is performed randomly in the population, e.g. universal random testing, then an unbiased estimator of the population prevalence would be 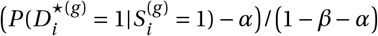.

### Extension to two tests

The above framework can be extended to handle allocation of two different types of tests with different cost and accuracy constrained by a fixed total budget. We suppose the first test option is cheap but has low sensitivity (e.g. rapid antigen testing) and the second test option is more expensive but has higher sensitivity (e.g. RT-PCR testing). Let 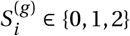 represent whether an individual is untested or given the first or second type of test, respectively. Following *Eq. 6*, the predicted numbers of positive tests of each type, denoted as 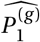 and 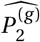 are:

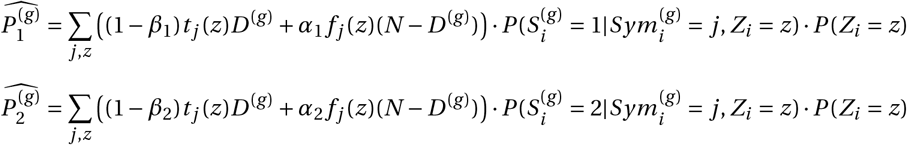

where *α*_1_, *α*_2_, *β*_1_, *β*_2_ are the false positive and negative rates corresponding to two tests. Suppose that the costs for one test of each type are *y*_1_ and *y*_2_. In the special case where our goal is to identify as many infected cases as possible (*w =* 1 in *Eq. 7*), we construct the following objective function for allocating two types of tests subject to a fixed total budget:

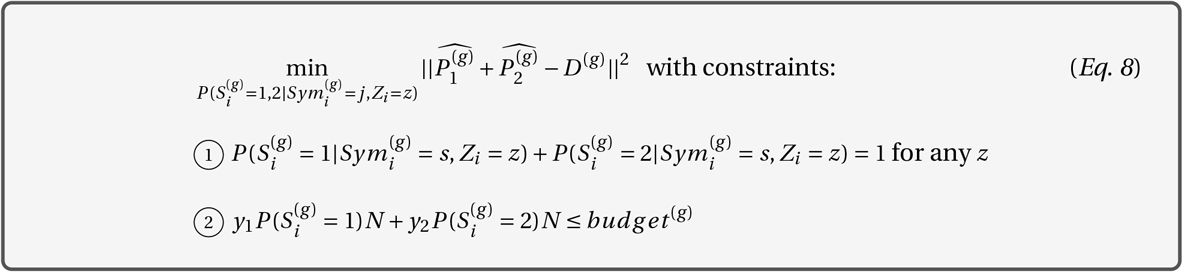

The first constraint guarantee that all the severe patients will be tested, and the second constraint ensures that the total spending does not exceed the budget. For the problem of assigning two tests, we do not consider to set *w =* 0 because it would always recommend to use the less accurate test for a lower test positive rate.

### Parameter estimates and setup

In the New York City example and all simulations, we estimate *t* _*j*_ (*z*) and *f* _*j*_ (*z*) using the publicly available historical data [29, 30]. For a given value of *t*_*a*_, the marginal probability for a truly infected person to be an asymptomatic carrier, the *marginal* probability of developing severe symptoms for an infected person is assumed to be roughly 1/4 of all COVID-19 symptomatic cases 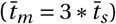, which is roughly the ratio of the number of hospitalizations to the cases until September 18, 2020 in the New York City [31]. For the uninfected person, the *marginal* probabilities of having severe 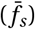 or mild 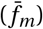 symptoms are set to 2.3 *×* 10^*−*5^ and 1.6 *×* 10^*−*4^, respectively, using the data from the New York State Department of Health 2019-2020 Flu Monitoring Archives [29]. In obtaining these estimates, hospitalized patients with flu-like symptoms were treated as severe cases, and the remaining laboratory-confirmed cases were treated as mild cases. We suppose *Z*_*i*_ represents age and is grouped into four categories: age 0 *−* 17, age 18 *−* 49, age 50 *−* 64 and age ≥ 65. Probabilities *t* _*j*_ (*z*) of having severe, mild and no symptoms for an infected person are approximated as 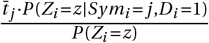. We estimated the age distribution within symptom categories from COVID-19 case counts and hospitalized counts by age from the New York City Department of Health and Mental Hygiene through September 18, 2020 [30]. Similarly, *f* _*j*_ (*z*) is estimated using data from the New York State Department of Health 2019-2020 Flu Monitoring Archives by age group [29]. **Table S3** provides estimates of *t* _*j*_ (*z*) and *f* _*j*_ (*z*) when 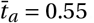.

When we evaluate the optimal strategy by simulation, we assume that 175, 000 people are truly infected at generation *g* (*D*^(*g*)^ *=* 175, 000), which mimic the situation of the peak of the pandemic. We suppose *T* ^(*g*)^, the number of available tests, to be either 50, 000 or 200, 000. These two scenarios correspond to the setting with limited testing resources and the setting with relatively sufficient testing resources. Due to the large number of infected cases in the population, the test strategy in this setting should be aimed at detecting cases, so the weight *w* in the objective function *Eq. 7* is set as 1. After the peak of the pandemic, we assume that the number of infected cases in the population deceases to 10, 000 (*D*^(*g*)^ *=* 10, 000) with 200,000 tests available. In this setting, our goal is for long-term surveillance to detect outbreaks, and we obtain optimal test allocation by setting *w =* 0 in *Eq. 7*. We define the outbreak threshold *c* to be 0.03 [32].

## Data Availability

Data used in this paper are all publicly available. Data sources are provided in the reference list.

## Acknowledgments

The work is supported by NSF DMS 1712933 and a pilot grant by the Michigan Institute of Data Science. The authors would like to thank Mike Kleinsasser for helping to build the R shiny application.

## Data Availability Statement

Data used in this paper are all publicly available. Links to the data are provided in the reference list.

## Author contributions

Jiacong Du, Lauren J. Beesley, and Bhramar Mukherjee conceptualized and formulated the mathematical framework. Jiacong Du drafted the manuscript, collected data, and wrote codes. Lauren J. Beesley built the R shiny app and co-drafted the manuscript. Seunggeun Lee, Xiang Zhou, Walter Dempsey, and Bhramar Mukherjee revised the manuscript. All participated in reading, editing and revising the initial draft of the manuscript.

