## Supplementary tables, figures and notes for "Optimal test allocation strategy during the COVID-19 pandemic and beyond"

### Supplementary materials for optimal test allocation strategy during the COVID-19 pandemic and beyond

Figure S1: Test positive rate for different underlying true number of cases. The surveillance test allocation strategy is given under the assumption that the true number of cases is 10,000 and the probability of being asymptomatic for an infected case is 0.55 ($\bar{t_a} = 0.55$). The red horizontal line is the threshold for disease outbreak.

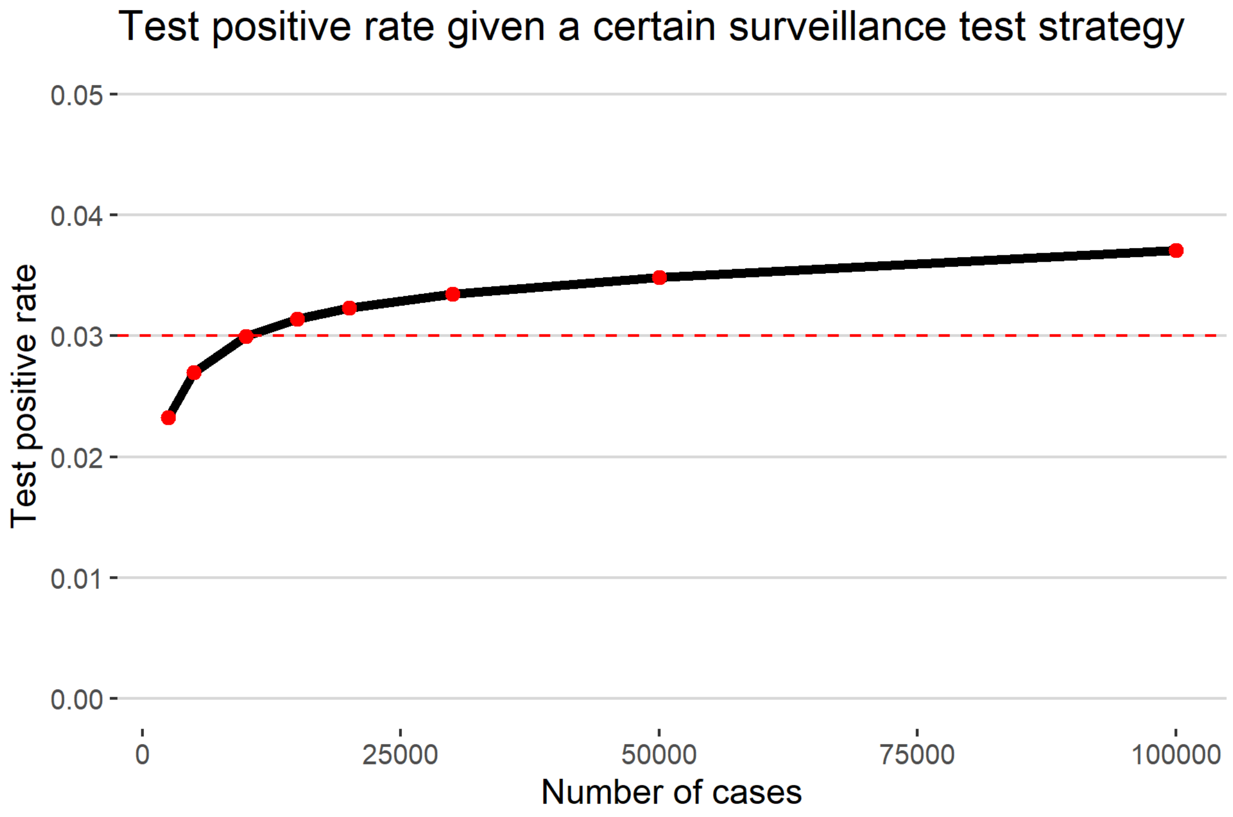

Table S1: Tests allocated to each symptom and age group from the proposed optimal strategy and other competing methods near the peak of the pandemic under the detecting mode, assuming that the number of true infected cases is 175,000 at generation $g$. The population size is 8 million. $\bar{t_a}$ is the probability for a person of being asymptomatic after being infected.

| Symptom | Severe | | | | Mild | | | | Asymptomatic | | | | Positive tests | |
|---|---|---|---|---|---|---|---|---|---|---|---|---|---|---|
| Age | 0-17 | 18-49 | 50-64 | 65+ | 0-17 | 18-49 | 50-64 | 65+ | 0-17 | 18-49 | 50-64 | 65+ | Number | % |
| **Limited number of tests as 50,000; $\bar{t_a}$=0.55** | | | | | | | | | | | | | | |
| Population | 236 | 3280 | 6605 | 10574 | 2974 | 27307 | 21456 | 8577 | 1916788 | 3409411 | 1651937 | 940848 | - | - |
| Detecting mode | 236 | 3280 | 6605 | 10574 | 0 | 0 | 21456 | 7846 | 0 | 0 | 0 | 0 | 34683 | 0.694 |
| Risk-based | 236 | 3280 | 6605 | 10574 | 0 | 0 | 20725 | 8577 | 0 | 0 | 0 | 0 | 34682 | 0.694 |
| Symptom-based | 236 | 3280 | 6605 | 10574 | 1445 | 13266 | 10423 | 4166 | 0 | 0 | 0 | 0 | 34456 | 0.689 |
| Severe-only | 236 | 3280 | 6605 | 10574 | 10 | 100 | 78 | 31 | 7039 | 12520 | 6066 | 3455 | 15049 | 0.301 |
| Universal random | 1 | 20 | 41 | 66 | 18 | 170 | 134 | 53 | 11979 | 21308 | 10324 | 5880 | 1252 | 0.025 |
| **Sufficient number of tests as 200,000; $\bar{t_a}$=0.55** | | | | | | | | | | | | | | |
| Population | 236 | 3280 | 6605 | 10574 | 2974 | 27307 | 21456 | 8577 | 1916788 | 3409411 | 1651937 | 940848 | - | - |
| Detecting mode | 236 | 3280 | 6605 | 10574 | 2974 | 27307 | 21456 | 8577 | 119001 | 0 | 0 | 0 | 58596 | 0.293 |
| Risk-based | 236 | 3280 | 6605 | 10574 | 2974 | 27307 | 21456 | 8577 | 0 | 0 | 0 | 118986 | 57087 | 0.285 |
| Symptom-based | 236 | 3280 | 6605 | 10574 | 2974 | 27307 | 21456 | 8577 | 28800 | 51228 | 24821 | 14136 | 57900 | 0.290 |
| Severe-only | 236 | 3280 | 6605 | 10574 | 66 | 613 | 482 | 192 | 43072 | 76612 | 37120 | 21141 | 18553 | 0.093 |
| Universal random | 5 | 82 | 165 | 264 | 74 | 682 | 536 | 214 | 47919 | 85235 | 41298 | 23521 | 5016 | 0.025 |

Table S1 (continued): Tests allocated to each symptom and age group from the proposed optimal strategy and other competing methods near the peak of the pandemic under the detecting mode, assuming that the number of true infected cases is 175,000 at generation $g$. The population size is 8 million. $\bar{t_a}$ is the probability for a person of being asymptomatic after being infected.

| Symptom | Severe | | | | Mild | | | | Asymptomatic | | | | Positive tests | |
|---|---|---|---|---|---|---|---|---|---|---|---|---|---|---|
| Age | 0-17 | 18-49 | 50-64 | 65+ | 0-17 | 18-49 | 50-64 | 65+ | 0-17 | 18-49 | 50-64 | 65+ | Number | % |
| **Limited number of tests as 50,000; $\bar{t_a}$=0.90** | | | | | | | | | | | | | | |
| Population | 66 | 754 | 1506 | 2221 | 1072 | 6405 | 4918 | 1980 | 1918860 | 3432839 | 1673574 | 955797 | - | - |
| Detecting mode | 66 | 754 | 1506 | 2221 | 1072 | 6405 | 4918 | 1980 | 31093 | 0 | 0 | 0 | 13034 | 0.261 |
| Risk-based | 66 | 754 | 1506 | 2221 | 1072 | 6405 | 4918 | 1980 | 0 | 0 | 0 | 31072 | 12952 | 0.259 |
| Symptom-based | 66 | 754 | 1506 | 2221 | 1072 | 6405 | 4918 | 1980 | 7470 | 13364 | 6515 | 3721 | 12994 | 0.260 |
| Severe-only | 66 | 754 | 1506 | 2221 | 6 | 36 | 27 | 11 | 10907 | 19514 | 9513 | 5433 | 4185 | 0.084 |
| Universal random | 0 | 4 | 9 | 13 | 6 | 40 | 30 | 12 | 11992 | 21455 | 10459 | 5973 | 1252 | 0.025 |
| **Sufficient number of tests as 200,000; $\bar{t_a}$=0.90** | | | | | | | | | | | | | | |
| Population | 66 | 754 | 1506 | 2221 | 1072 | 6405 | 4918 | 1980 | 1918860 | 3432839 | 1673574 | 955797 | - | - |
| Detecting mode | 66 | 754 | 1506 | 2221 | 1072 | 6405 | 4918 | 1980 | 181089 | 0 | 0 | 0 | 16768 | 0.084 |
| Risk-based | 66 | 754 | 1506 | 2221 | 1072 | 6405 | 4918 | 1980 | 0 | 0 | 0 | 181072 | 16289 | 0.081 |
| Symptom-based | 66 | 754 | 1506 | 2221 | 1072 | 6405 | 4918 | 1980 | 43534 | 77883 | 37969 | 21684 | 16537 | 0.083 |
| Severe-only | 66 | 754 | 1506 | 2221 | 26 | 156 | 120 | 48 | 46906 | 83916 | 40910 | 23364 | 7894 | 0.039 |
| Universal random | 1 | 18 | 37 | 55 | 26 | 160 | 122 | 49 | 47971 | 85820 | 41839 | 23894 | 5015 | 0.025 |

Table S2: Tests allocated to each symptom and age group from the proposed optimal strategy and other competing methods after the peak of the pandemic under the surveillance mode, assuming that the number of true infected cases is 10,000 at generation $g$. The population size is 8 million. $\bar{t}_a = 0.55$ is the probability for a person of being asymptomatic after being infected. The total number of available tests is 200,000. The actually number of used tests may not be added up to the exact number of available tests due to rounding of numbers.

| Symptom | Severe | | | | Mild | | | | Asymptomatic | | | | Used tests | Positive tests | |
|---|---|---|---|---|---|---|---|---|---|---|---|---|---|---|---|
| Age | 0-17 | 18-49 | 50-64 | 65+ | 0-17 | 18-49 | 50-64 | 65+ | 0-17 | 18-49 | 50-64 | 65+ | | Number | % |
| Population | 30 | 218 | 425 | 628 | 680 | 1978 | 1412 | 582 | 1919288 | 3437802 | 1678161 | 958789 | | - | - |
| Surveillance mode | 30 | 218 | 425 | 628 | 342 | 1088 | 763 | 300 | 19327 | 2521 | 24479 | 52956 | 103077 | 3090 | 0.030 |
| Risk-based | 30 | 218 | 425 | 628 | 680 | 1978 | 1412 | 582 | 0 | 0 | 0 | 194042 | 199995 | 5125 | 0.026 |
| Symptom-based | 30 | 218 | 425 | 628 | 680 | 1978 | 1412 | 582 | 46587 | 83446 | 40734 | 23273 | 199993 | 5194 | 0.026 |
| Severe-only | 30 | 218 | 425 | 628 | 16 | 49 | 35 | 14 | 47677 | 85398 | 41687 | 23817 | 199994 | 2926 | 0.015 |
| Universal random | 0 | 5 | 10 | 15 | 17 | 49 | 35 | 14 | 47982 | 85945 | 41954 | 23969 | 199995 | 2170 | 0.011 |
| Detecting mode | 30 | 218 | 425 | 628 | 680 | 1978 | 1412 | 582 | 194049 | 0 | 0 | 0 | 200002 | 5261 | 0.026 |

Figure S2: The number of each type of test allocated to each symptomatic group. The total budget for testing is 1 million. The price of a single antigen test and RT-PCR test are \$ 5 and \$ 135, respectively. The number of infected cases is assumed to be 10,000 in a region of 8 million people.

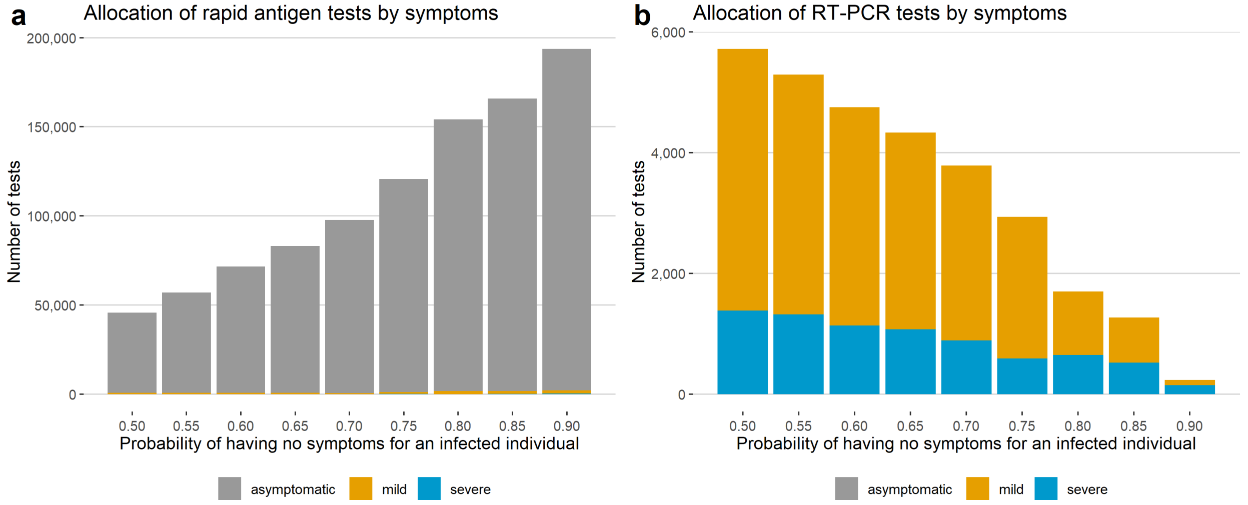


#### Relationship between test positive rate and disease prevalence

Let $r$ be the ratio of selection probability in the infected population versus the uninfected population; that is, $r = \frac{P(S_i^{(g)}=1|D_i^{(g)}=1)}{P(S_i^{(g)}=1|D_i^{(g)}=0)}$. Let $\beta, \alpha, c$ be the test false negative rate, false positive rate and the outbreak threshold test positive rate. If $r > 1$, $1-\beta > c$ and $1-\beta > \alpha$, then the disease prevalence $P(D_i^{(g)} = 1) < \frac{c-\alpha}{(1-\beta-c)r+(c-\alpha)}$.

*Proof:* The derivation is straightforward from:

$$P(D_i^{\star(g)} = 1|S_i^{(g)} = 1) = \frac{(1-\beta)rP(D_i^{(g)} = 1) + \alpha(1-P(D_i^{(g)} = 1))}{rP(D_i^{(g)} = 1) + (1-P(D_i^{(g)} = 1))} < c$$

| age | $t_s(z)$ | $t_m(z)$ | $t_a(z)$ | $f_s(z)$ | $f_m(z)$ | $f_a(z)$ |
|---|---|---|---|---|---|---|
| 0-17 | 5.816E-02 | 5.186E-03 | 9.367E-01 | 9.619E-06 | 2.821E-04 | 9.997E-01 |
| 18-44 | 3.571E-01 | 4.307E-02 | 5.998E-01 | 9.642E-06 | 1.287E-04 | 9.999E-01 |
| 45-64 | 5.786E-01 | 1.783E-01 | 2.431E-01 | 3.024E-05 | 1.178E-04 | 9.999E-01 |
| 65+ | 4.041E-01 | 4.608E-01 | 4.608E-01 | 7.907E-05 | 1.017E-04 | 9.998E-01 |

Table S3: Estimated probabilities of having severe, mild and no symptoms for the infected individual when the marginal probability of being asymptomatic after infected is $0.55$ ($\bar{t_a} = 0.55$), and the probability of developing severe symptoms is $1/3$ of developing mild symptoms ($\bar{t_s} = 1/3\bar{t_m}$). $t_j(z)$ is the probability of developing symptom $j$ conditional on a person being infected in age group z. Estimates are obtained using the distribution of age in the hospitalized patients and overall cases in New York City. $f_j(z)$ is the probability of developing symptom $j$ in age group $z$ for the uninfected person. Estimates are obtained by using the number of flu hospitalizations and cases from New York State Department of Health 2019-2020 Flu Monitoring Archives.

Table S4: Testing information for some universities around the week ending at Novermber 14, 2020.

| University | # tests | # positive tests | test positive % |
|---|---|---|---|
| Ohio State University | 14856 | 378 | 0.03 |
| Michigan State University | 1398 | 164 | 0.12 |
| Indiana University | 12130 | 207 | 0.02 |
| Purdue University | 6684 | 437 | 0.07 |
| University of Michigan | 8319 | 152 | 0.02 |
| University of Minnesota | 592 | 81 | 0.14 |
| University of Wisconsin-Madison | 12466 | 458 | 0.04 |
| University of Illinois-Urbana-Champaign | 64239 | 290 | 0.00 |
| University of Nebraska | 1565 | 157 | 0.10 |
| Pennsylvania State University | 5552 | 264 | 0.05 |
| University of Maryland | 4135 | 50 | 0.01 |
| Rutgers University | 4652 | 93 | 0.02 |
